# Trends of SARS-Cov-2 infection in 67 countries: Role of climate zone, temperature, humidity and curve behavior of cumulative frequency on duplication time

**DOI:** 10.1101/2020.04.18.20070920

**Authors:** Jaime Berumen, Max Schmulson, Guadalupe Guerrero, Elizabeth Barrera, Jorge Larriva-Sahd, Gustavo Olaiz, Rebeca Garcia-Leyva, Rosa María Wong Chew, Miguel Betancourt-Cravioto, Héctor Gallardo, Germán Fajardo-Dolci, Roberto Tapia-Conyer

**Affiliations:** Unidad de Medicina Experimental, Facultad de Medicina, Universidad Nacional Autónoma de México, Mexico City, Mexico; Hospital General de México, Dr. Eduardo Liceaga, Mexico City, Mexico; Instituto de Neurobiología, Universidad Nacional Autónoma de México, Campus Juriquilla, Querétaro Mexico; Centro de Investigación en políticas, población y salud, Facultad de Medicina, Universidad Nacional Autónoma de Mexico, Mexico City, Mexico; División de Investigación, Facultad de Medicina, Universidad Nacional Autónoma de México, Mexico City, Mexico; Fundación Carlos Slim, Mexico City, Mexico; Facultad de Medicina, Universidad Nacional Autónoma de México, Mexico City, Mexico

**Keywords:** COVID-19, doubling time, temperature, humidity, curve behavior, cumulative frequency, date, temperate, tropical, subtropical

## Abstract

**Objective:** To analyze the role of temperature, humidity, date of first case diagnosed (DFC) and the behavior of the growth-curve of cumulative frequency (CF) [number of days to rise (DCS) and reach the first 100 cases (D100), and the difference between them (ΔDD)] with the doubling time (Td) of Covid-19 cases in 67 countries grouped by climate zone.

**Design:** Retrospective incident case study.

**Setting:** WHO based register of cumulative incidence of Covid-19 cases.

**Participants:** 1,706,914 subjects diagnosed between 12-29-2019 and 4-15-2020.

**Exposures:** SARS-Cov-2 virus, ambient humidity, temperature and climate areas (temperate, tropical/subtropical).

**Main outcome measures:** Comparison of DCS, D100, ΔDD, DFC, humidity, temperature, Td for the first (Td10) and second (Td20) ten days of the CF growth-curve between countries according to climate zone, and identification of factors involved in Td, as well as predictors of CF using lineal regression models.

**Results:** Td10 and Td20 were ≥3 days longer in tropical/subtropical vs. temperate areas (2.8±1.2 vs. 5.7±3.4; p=1.41E-05 and 4.6±1.8 vs. 8.6±4.2; p=9.7E-05, respectively). The factors involved in Td10 (DFC and ΔDD) were different than those in Td20 (Td10 and climate areas). After D100, the fastest growth-curves during the first 10 days, were associated with Td10<2 and Td10<3 in temperate and tropical/subtropical countries, respectively. The fold change Td20/Td10 >2 was associated with earlier flattening of the growth-curve. In multivariate models, Td10, DFC and ambient temperature were negatively related with CF and explained 44.7% (r^2^ = 0.447) of CF variability at day 20 of the growth-curve, while Td20 and DFC were negatively related with CF and explained 63.8% (r^2^ = 0.638) of CF variability towards day 30 of the growth-curve.

**Conclusions:** The larger Td in tropical/subtropical countries is positively related to DFC and temperature. Td and environmental factors explain 64% of CF variability in the best of cases. Therefore, other factors, such as pandemic containment measures, would explain the remaining variability.

## Introduction

In December 2019, a new epidemic of severe acute respiratory syndrome (SARS) virus broke out in China, in the city of Wuhan, province of Hubei. It was initially termed novel Coronavirus 2019, (nCoV-2019) later called the severe acute respiratory syndrome-coronavirus-2 (SARS-CoV-2), and after, the World Health Organization (WHO) denominated it coronavirus disease-2019 (COVID-19).^1-3^ It was called SARS-CoV-2 considering that it was the second SARS epidemic after the first one in 2002, and both had in common that they originated in China.^4^ The current SARS-Cov-2 spread from China to other Asian countries including Thailand, South Korea and Japan and then Australia. Subsequently, new cases appeared in Europe, the United States, Canada and Iran, and by the end of February 2020 there were already cases in Brazil, Mexico, Greece and Norway among others; and at the beginning of March, the first cases appeared in Argentina. On March 13, there was an increase in the number of cases in western European countries and the illness also emerged in African countries such as Sudan, Ethiopia, Kenya, and Guinea. On March 22, there were new cases in countries from all continents with increasing numbers in the United States and South America.^5^ By April 11^th^, 2020, a total of 1,254,464 cases had been confirmed worldwide, with 68,184 (5.4%) deaths.^6^ According to the above reports, the infection rate among the total population was much higher in Europe than in the Americas. In Europe for example, as of April 5^th^, 2020, the infection rate varied from 0.06% in Sweden to 0.27% in Spain. However, there were some European countries with lower infection rates including 0.07% in the United Kingdom albeit a high population density of 281 inhabitants per Km^2^, or 0.10% in the Netherlands despite a much higher population density of 508 inhabitants per Km^2^ or 0.11% in Germany with a 240 inhabitant density per Km^2^.^6^ In contrast, in the Americas, up to the same date in April, the United States reported the highest number of cases with a total of 327,920. However, this only comprised 0.01% of its total population, and Canada had a 0.04% infectious rate. The data from these North American countries, were low, but still 10 times more frequent than in the rest of the countries of Central and South America, with rates ranging from 0.04% in Panama, 0.001% in Mexico, 0.002% in Colombia and 0.0003% in Guatemala.^6^ The above described infectious rates, decreasing from Europe to North America and South America, is in agreement with the time frame of the spreading path that this pandemic has followed.^5^

However, the much lower infection rates in South America, compared to the USA and Canada as well as that in Europe, may also reflect an underestimation on the illness, although other variables including climatologic differences deserve to be studied. First, the cold winter season and severe drought were both present in China during the first SARS 2002 and the current COVID-19 epidemics.^4^ Second, the combination of cold and dry environment seem to be more adequate than cold weather by itself for viral transmission.^7^ Even more, cold temperature and lower humidity reduce both ciliary movement and the mucous secretion as defensive mechanism to remove particles such as infectious agents, and alters the integrity of the nose-mucosal epithelium. Overall, cold and dry environment may enhance the susceptibility to COVID-19 infections.^4, 8^

In addition, cold season and low temperatures cause stress to the human organism impairing the immune system, increasing norepinephrine and cortisol levels, producing lymphocytosis and decreasing the lymphoproliferative responses, and TH1 cytokine levels and salivary IgA hemostasis.^8^ These mechanisms may be predisposing factors to acquire viral infections in winter.^8^

Although in Europe and North America, including the USA and Canada, cold weather remains even as spring season has begun, in the northern countries of Latin America such as Mexico, the predominant climate is warm humid weather. And in more southern countries such as Argentina, it is autumn with template weather. Thus, it is plausible that the higher temperatures and humidity in these countries, explain the much lower infectious rates of COVID-19 when compared to the Northern hemisphere. ^9^ Also, viral infections have cyclic patterns such as the annual flu or the human respiratory syncytial virus epidemic in which hosts are more susceptible during the winter months.^10^ However, not all viruses have the same seasonality, and some spread during the summer season (e.g. Enteroviruses), other have a spring/fall cycle (e.g. Rhinovirus), or all year round (e.g. Adenovirus).^11^ Also, transmission can occur in different weather patterns.^12, 13^ which is especially true for tropical regions in which there is no clear seasonal pattern.^14^ When a new virus such as the SARS CoV2 arise with no immunity for humans, and with a great capacity to disseminate, the seasonality might or might not be a determinant factor for its spread.^15^

Therefore we aimed at analyzing the role of climate areas, outside temperature, humidity, the number of days it took for the spreading curve to raise or elevate (DCS) and to reach the first 100 cases (D100) and the difference of the latter ones (ΔDD) with the duplication speed of infected patients, in 67 countries around six continents. We hypothesized that the elevation of the curve, the time to reach the initial 100 cases, as well as the time for duplicating the number of cases, should be faster in number of days in countries within lower temperate areas when compared to those in tropical or subtropical ones.

## Material and methods

### Study population

The study populations are the daily confirmed newly diagnosed cases of COVID-19 officially reported by the WHO from 67 countries, 18 located in temperate or cold areas, and 49 in tropical or subtropical regions from December 29, 2019 to April 15, 2020. The population data was collected from the reports released on the official websites of the World Health Organization about Covid-2019.^16^ The cumulative frequency (CF) and the date when the first case was diagnosed in each country were obtained. Thus, no ethical review was required.

### Average, minimum and maximum temperatures

The average temperature and relative humidity were collected from Time and Date AS database during the months of January, February and March for each of the 67 countries analyzed.^17^

### Calculation of doubling time and the parameters of the growth curve of CF of Covid-19 cases

The CF of Covid-19 cases of each country was plotted in Excel and the exponential equation was obtained. The days of the curve rise (DCS) and of reaching the first 100 cases (D100), as well as the difference between these two days (ΔDD) were graphically identified with the WHO data. The duplication time (Td) of the number of positives was obtained from the slope (λ) of the exponential graph (N=Noe^λt^) as follows: Td=ln(2)/λ.^18^ It was calculated for the first (Td10), second (Td20) and third (Td30) 10-day periods of the CF curve, as well as for the entire 30-day period (TdT), starting from day D100.

### Statistical analysis

Descriptive analysis was performed. Numerical variables were described with medians and interquartile range (IQR) or means and standard deviations. The variables were compared between the groups (temperate vs. tropical/subtropical zones). The significance of differences between the groups was assessed with the Mann-Whitney U-test or the t-test. The Pearson’s correlation was performed for some numerical variables. The association of significant variables with Td was explored using univariate (ULR) and multivariate (MLR) linear regression models. Finally, we built models to predict the CF of Covid-19 cases expected to happen on day 20 and 30 of the pandemic growth curve with the Td and the rest of variables using ULR and MLR models. To enter the date of the first Covid-19 case diagnosed and the climate zone in the linear regression models the nominal data were transformed into numeric values, as follows: 1) the dates of the first Covid-19 cases diagnosed in the 67 countries were sorted in ascending order, and numerical values in ascending order, starting with the number 1, were assigned to each date, 2) for the climate zones, the values of 1 and 2 were assigned to temperate and tropical/subtropical zones, respectively.

The association was expressed as the β coefficient and 95% confidence interval (CI), and the contribution to the variability of Td or CF was expressed as adjusted r^2^. Confounders were identified using a theoretical strategy based on a backstep, stepwise MLR model and the change-in-estimate criterion. Variables with p < 0.20 in the univariate analysis were considered for entry in the multivariate model. Confounders were defined as those variables for which the percentage difference between the values of the regression β and the adjusted and non-adjusted variables in the stepwise multivariate model were higher than 10% (p>0.1). Therefore, the total variability, the contribution of each factor, and interaction between the factors on Td or CF was calculated using this MLR models. The factors and interactions were included in the model in one block. A post hoc power analysis was performed for each linear regression model using the software G * Power 3.1.9.2, considering the sample size, the β and an α = 0.05. In addition, for MLR models, the value of the total r^2^ obtained at the end of the model was introduced for power calculation. All statistical tests were two-sided, and differences were considered significant when p < 0.05. The statistical analyses were conducted using SPSS version 20 software (SPSS Inc., Chicago, IL, USA).

## Results

### Analysis of the initial phase of the growth curve

Figure 1 shows how the CF grew from the day the first case of Covid-19 was diagnosed in 67 countries. The day the curve started to ascend (DCS) and the slope of the curves were observed. There is a wide variation between countries, but similar patterns are generally observed across continents or climatic regions. Unexpectedly, in countries located in temperate areas, such as most of the European countries, USA, Canada, Japan and Korea, the rise of the curve began late, 20 days after the first case was diagnosed. Conversely, in most countries located in tropical or subtropical areas such as those in the Middle East, Africa, Mexico, Central and South America, Asia and Oceania, the rise of the curves began much earlier. However, in this group there are notable exceptions such as New Zealand, Australia, Egypt and India, where the curve started belatedly. In fact, the median number of days it took for the curves to start to ascend was much higher in temperate countries than in tropical/subtropical ones [median (IQR) 29 (8-32) vs. 12 (8-16), p=0.015; Man-Whitney U-test; Figure 2 panel A). On the other hand, the slope of the curves was much higher in temperate than in tropical/subtropical countries (Figure 1). An indirect way to evaluate the slope, is to measure the number of days it takes for the curve to reach 100 cases, once the ascend begins. Thus, we identified the number of days it took for the curves to reach 100 cases (D100) from the diagnosis of the first case and subtracted the DCS to obtain the difference in days between the two points (ΔDD). While the D100 is much higher [median (IQR) 32 (12-36) vs. 20 (14-25); p=0.108, Man-Whitney U-test; Figure 2 panel B], the ΔDD is much lower in temperate countries [median (IQR) 3 (2-5) vs 7 (6-9); p=0.002, Man-Whitney U-test; Figure 2 panel C] than in tropical or subtropical areas. This suggests that the rate for doubling the number of positive cases is higher in temperate countries.

**Figure 1.**
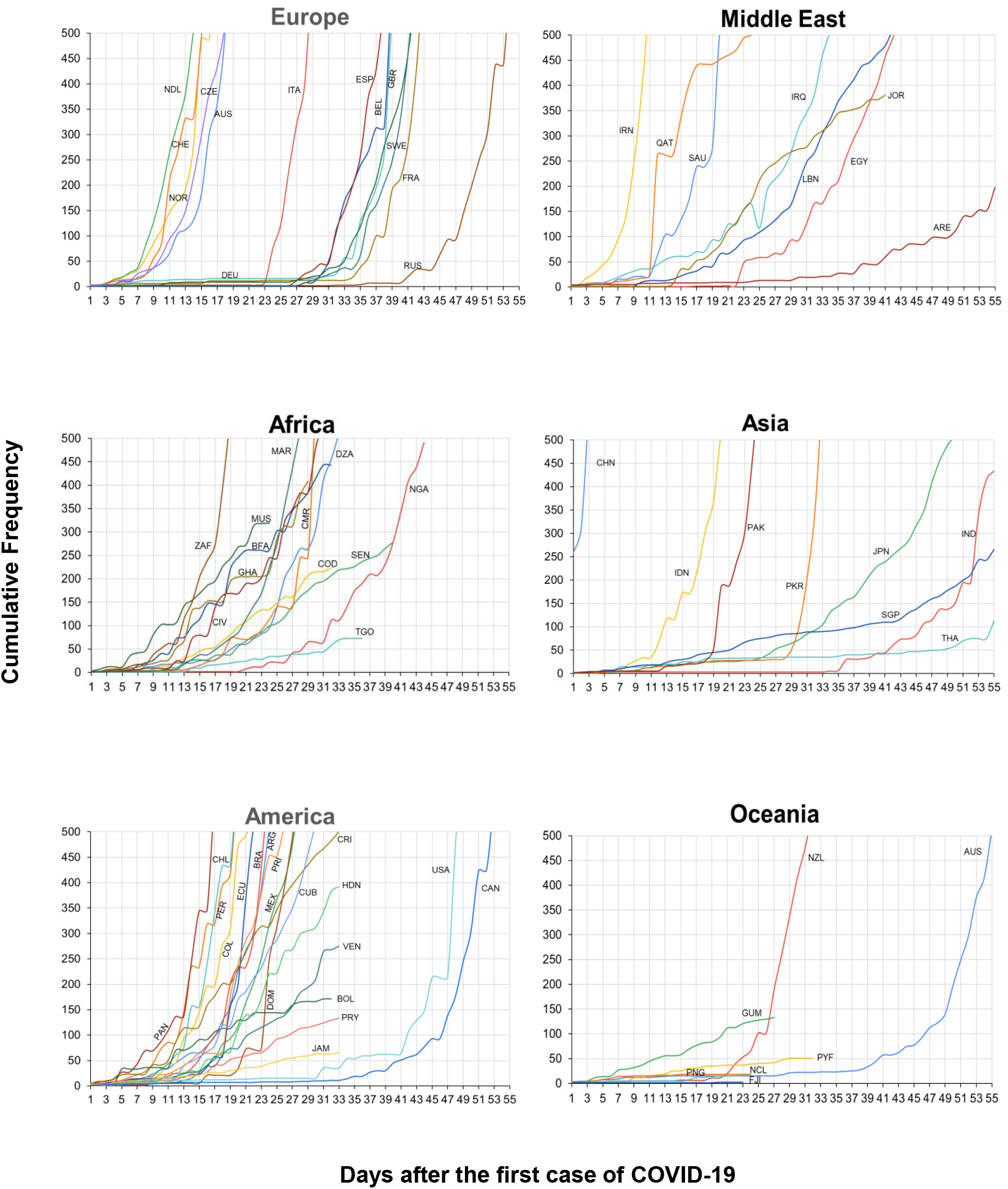
Days when the curves started to climb (DCS) and reach the first 100 (D100) Covid-19 cases. The curves show the cumulative frequency (CF) starting on the day when the first case of COVID-19 was diagnosed in each country. Based on these graphs we selected the days when the curves begin to climb and reach the first 100 cases. Abbreviations of each country: CHN: China, THA: Thailand, JPN: Japan, PKR: Republic of Korea, USA: USA, SGP: Singapore, AUS: Australia, FRA: France, CAN: Canada, DEU: Germany, ARE: United Arab Emirates, IND: India, ITA: Italy, RUS: Russian Federation, SWE: Sweden, GBR: United Kingdom, ESP: Spain, SAU: Saudi Arabia, BEL: Belgium, EGY: Egypt, IRN: Iran, LBN: Lebanon, IRQ: Iraq, CHE: Switzerland, AUT: Austria, BRA: Brazil, NOR: Norway, DZA: Algeria, PAK: Pakistan, NDL: Netherlands, NGA: Nigeria, NZL: New Zealand, MEX: Mexico, ECU: Ecuador, QAT: Qatar, CZE: Czech Republic, DOM: Dominican Republic, IDN: Indonesia, JOR: Jordan, MAR: Morocco, SEN: Senegal, CHL: Chile, ARG: Argentina, CMR: Cameroon, ZAF: South Africa, PER: Peru, COL: Colombia, CRI: Costa Rica, PAN: Panama, BOL: Bolivia, BFA : Burkina Faso, COD: Republic of Congo, HDN: Honduras, CIV: Ivory Coast, CUB: Cuba, PRI: Puerto Rico, VEN: Venezuela, GHA: Ghana, MUS: Mauritius, TGO: Togo, PRY: Paraguay, JAM: Jamaica, PNG: Papua New Guinea, NCL: New Caledonian, GUM: Guam, PYF: French Polynesia, FJI: Fiji.

**Figure 2.**
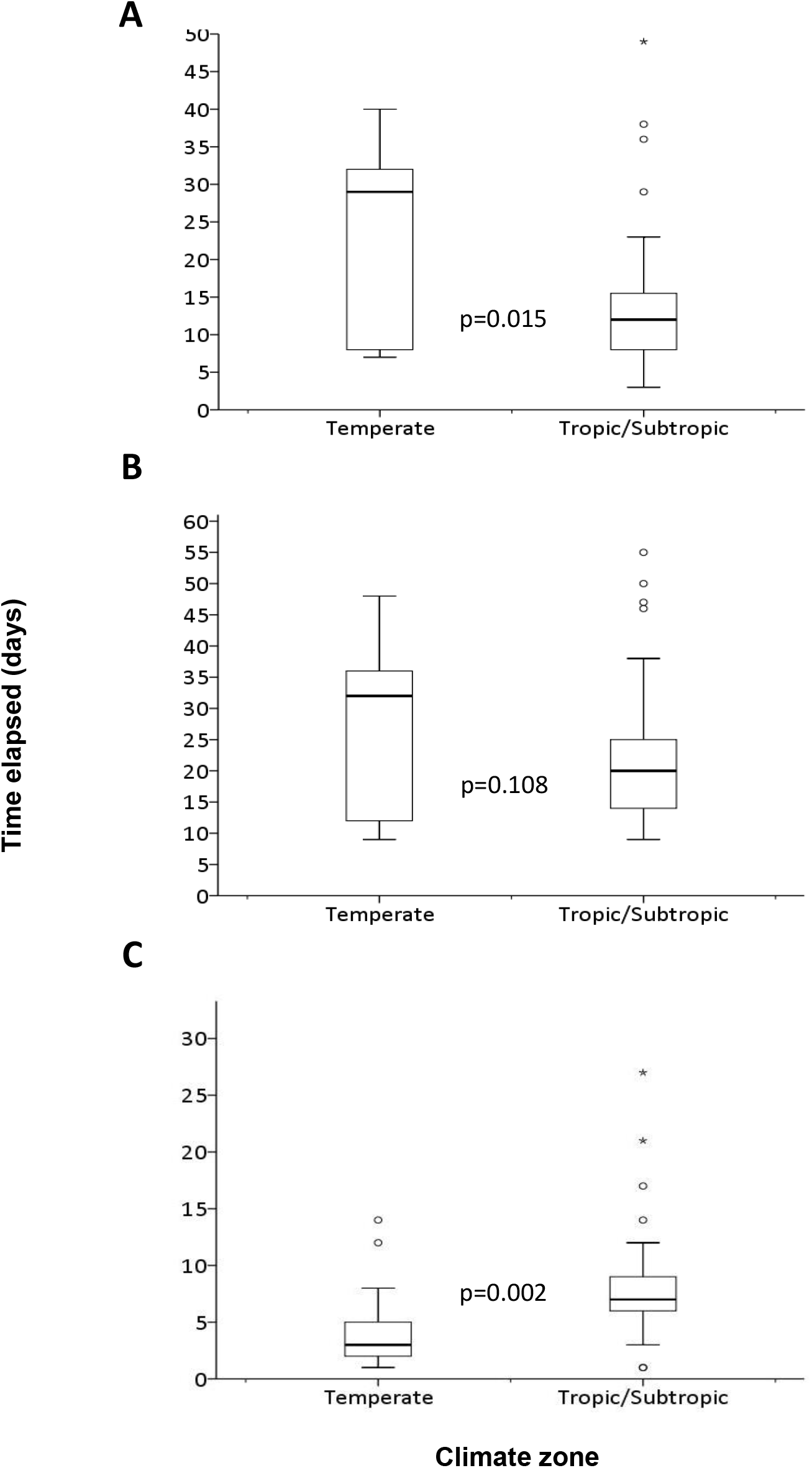
Time elapsed in days from the first case of Covid-19 diagnosed to the DCS and the day when 100 cases was reached (D100). Panel A shows a box plot distribution of DCS, panel B the distribution of D100 and panel C the distribution of these point-differences (ΔDD). The data included the analysis of 67 countries. Medians were compared between the groups by the Man-Whitney U-test.

Interestingly, we found that the date in which the first case of Covid-19 was diagnosed had a linear negative correlation with the DCS (r= -0.75; p= 1.3E-29, Pearson correlation; Figure 3 panel A) and the D100 (r= -0.71; p= 4.4E-19, Pearson correlation; Figure 3 panel B); that is, the later the first case was diagnosed, the faster the DCS and D100 days were reached; and vice versa, in countries where the first Covid-19 case was diagnosed very early in the year, for example in January, the DCS and D100 were longer (Figure 3 panels A and B, respectively). Although there is no significant linear correlation between dates in which the first case and ΔDD, because of data dispersion, the separation of the values of temperate (red circles) and tropical/subtropical countries (blue circles) is clearly observed (Figure 3 panel C). These correlations could be related to temperature and relative humidity, since the ascending dates from January to March correlated with those two variables (r= 0.658, p= 2.7E-26, and r= 0.233, p= 0.001, Pearson correlation). However, in the analysis of the data, only the DCS day correlated negatively with the temperature (r= -0.345; p= 5.4E-7, Pearson correlation).

**Figure 3.**
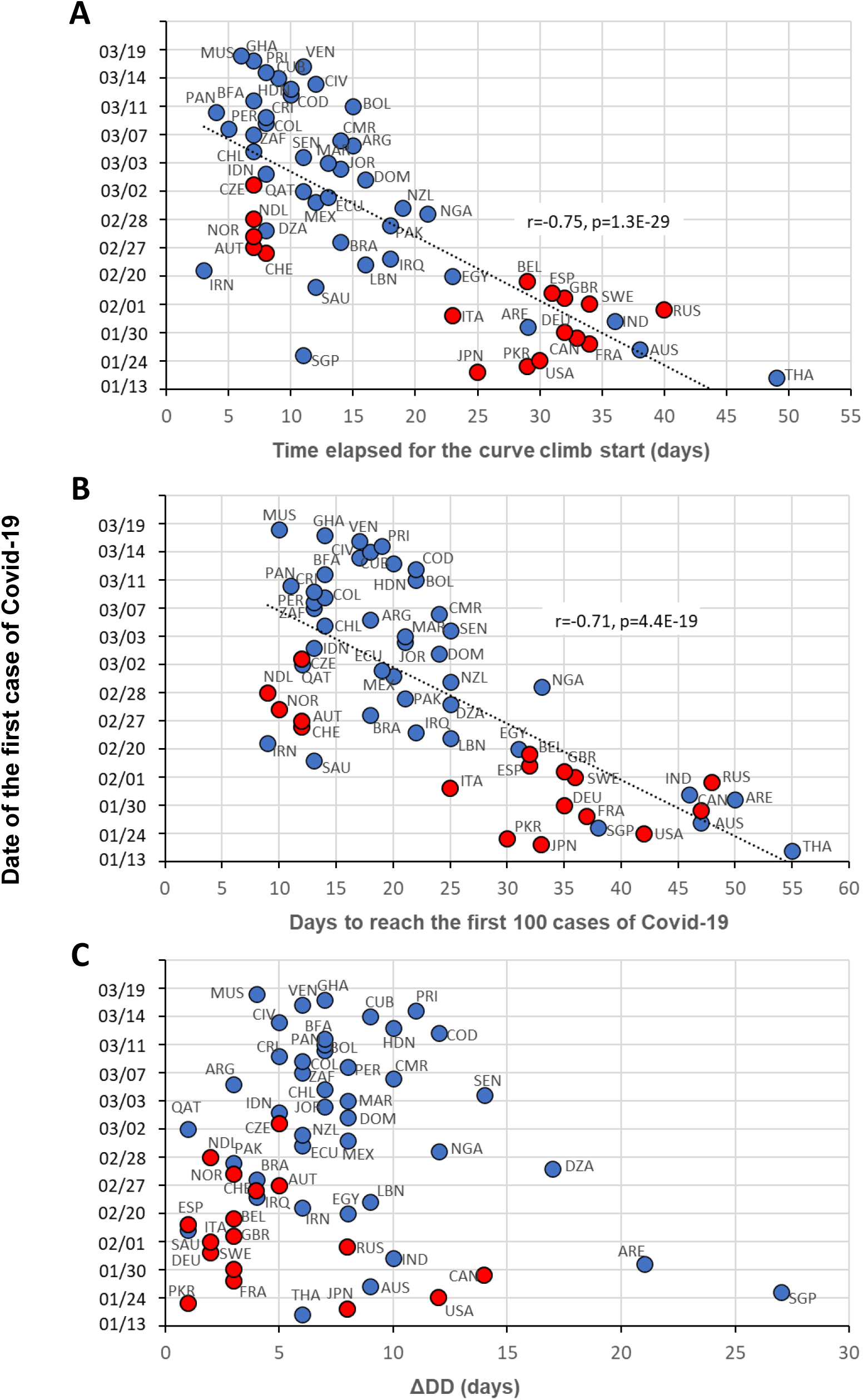
Correlation between the date of the first COVID-19 case diagnosed and DCS, D100 and ΔDD. The blue circles indicate the countries located in tropical/subtropical zones and red circles the countries located in temperate zones. Correlation coefficients and p-values were calculated with the Pearson’s correlation test. Abbreviations of each country are described in Figure 1-legend.

### Analysis of the growth curve in the first 30 days after the D100 day: calculation and analysis of the doubling time

The cumulative frequency, converted to log_10_, was plotted against the number of days of evolution of the epidemic in each country from D100 (Figure 4). In addition, doubling times of 1 to 4 days were calculated and included in the graph to locate the growth curve of each country between these intervals (black dotted lines in Figure 4). Changes in the trends of the accumulated frequencies over time are represented. For example, Figure 4 shows how the epidemic quickly grew in China, Korea and Iran during the first 10 days closer to Td= 1 day (dotted line, Figure 4), then the curves start to lie down after the 10th and 20th days of the evolution of the epidemic. Something similar was seen in the European countries. To compare the growth curves in more detail, the doubling time was calculated for the first (Td10), second (Td20), and third (Td30) 10 days, and for the entire 30-day period (TdT) in each country. The TdT was lower in temperate areas when compared with tropical/subtropical areas [4.2 ± 1.5 vs. 6.5 ± 3.1; p= 3.08E-04]; this difference became greater when comparing the Td10 (2.8 ± 1.2 vs. 5.7 ± 3.4; p= 1.41E-05) and the Td20 (4.6 ± 1.8 vs. 8.6 ± 4.2; p= 9.7E-05) (all with t-test). This clearly indicates that Td is lower in temperate countries.

**Figure 4.**
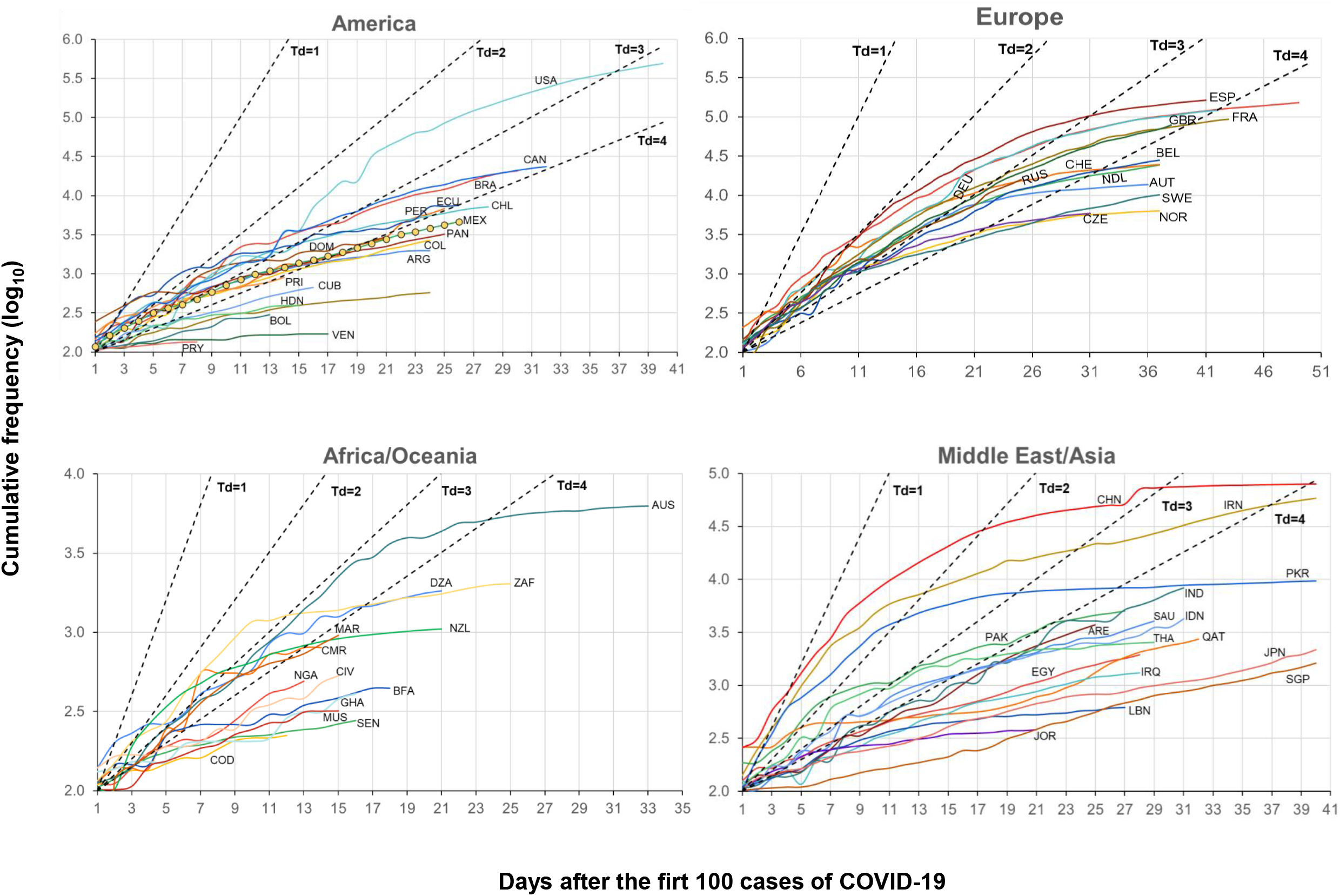
Trends of accumulative frequency in log10 starting the day when 100 COVID-19 cases were reached. The accumulated frequency (CF) is plotted against the number of days when 100 cases of Covid-19 were diagnosed in each country. The dashed black lines indicated the doubling times (Td) in days, that is the number needed to duplicate the number of infected people from one (Td=1) to 4 (Td=4) days. Abbreviations of each country are described in Figure 1-legend.

In addition, Figure 5 panels A and B depict the individual Td10, Td20 and Td30 from several temperate and tropical or subtropical countries. Countries that had a very rapid increase in the growth curve such as Italy, Spain, China, Korea and Iran (see Figure 4), have a Td10 very close to or below 2. It is important to point out that Td20 was higher than Td10 in all countries except the USA (Figure 5 panel A) and Singapore (Figure 5 panel B), which indicates that the rate for doubling the number of infected people was lower in most countries during days 11 to 20 of the curve. In the case of the USA, it is possible that the infection duplication rate grew during that time period. In fact, the Td20 falls below 2, and as seen in Figure 4, the slope of the curve increases from day 7 and the curve ascends almost as a straight line between Td= 2 and Td= 3. Something similar happened with Singapore (Figure 4), however, the curve remained below the Td= 4 line, which indicates that the Td > 4 days. In temperate countries, where the value of Td20 is two times that of Td10 [Fold Change (FC_Td20/Td10_)], such as Korea (FC_Td20/Td10_ = 4.5), Sweden (FC_Td20/Td10_= 2.5), Norway (FC_Td20/Td10_= 2.6), or that have a close value as in the case of the Czech Republic (FC_Td20/Td10_= 1.9; Figure 5), coincides with the early flattening of the curves, growing towards the area of Td= 4 or to a larger Td (Figure 4). It seems clear that the increase of the Td20, more than Td30, is essential for the early flattening of the curve. Examples of this include Austria (FC_Td20/Td10_= 1.6) and Switzerland (FCT_d20/Td10_= 1.5) that have a high Td30 but did not have a FCd_20/Td10_ > 2, and the curves flattened later. This is even more difficult for Spain, France and Italy that started with a very low Td10 and FCTd_20/Td10_= 1.7.

**Figure 5.**
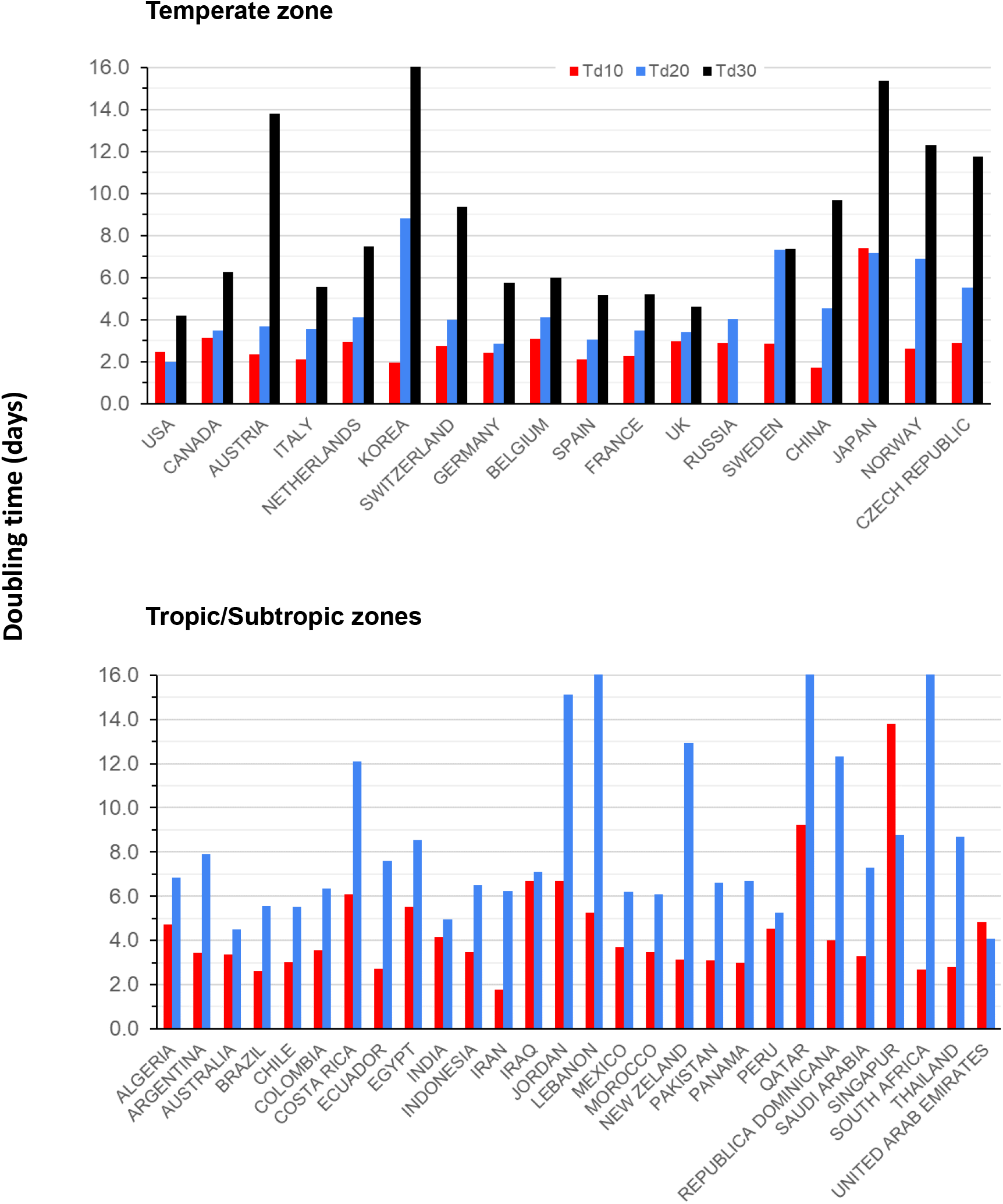
Doubling times (Td) in countries within temperate and tropical/subtropical zones. The plots show the Td from the first (blue), second (red) and third (black) 10 days of the cumulative frequency curves. Panel A shows countries from temperate zones and panel B countries from tropical/subtropical zones.

In tropical and subtropical areas, in addition to Iran that had a Td10= 1.8, countries that had a Td10 ≤ 3, such as Brazil, Chile, Ecuador and South Africa, had fast-growing curves during the first 10 days. However, most of them, except for Chile, had a FCTd_20/Td10_ ≥ 2.

In conclusion, it seems clear that the Td10 value is closely related to the initial momentum of the curve growth, with a cut-off point of Td10= 2 and Td10= 3, for temperate and tropical/subtropical countries, respectively, above which the growth of the curve is more manageable. The other important time-point is Td20, and when its value is ≥ 2 times that of Td10, there is a trend for flattening the curve earlier than when it is lower than 2.

### Identification of variables that could explain the doubling time using linear regression models

The relation of each variable (DCS, D100, ΔDD, first case date, average relative humidity, average temperature, climate zone) with Td10 and Td20 was investigated in ULR models. Variables that had a significance of p< 0.2 were selected to be introduced in multivariate models, one for the Td10 and one for the Td20, and variables were selected in the final models when they remained in the MLR analysis with a p< 0.1. In addition, Td10 was included in the Td20 model (Table 1). Five of these variables were individually associated with the Td10, four showed a positive association (ΔDD, date of first case diagnosed, average temperature and climate zone), while DCS had a negative association. The average relative humidity and the D100 day were not associated with the Td10. The average temperature (β= 0.17 95%CI: 0.092 to 0.247, p= 4.7E-05) and the date of the first case (β=0.079 95%CI: 0.037 to 0.121, p= 4E-04) were the variables that most influenced the value of Td10; in fact, they also showed the highest values of r^2^ (0.237 and 0.182, respectively), which multiplied by 100, indicated the percentage of Td10 variability explained by that variable. However, in the MLR analysis, the only variables that remained in the model were the date of the first case diagnosed (β= 0.077 95%CI: 0.037 to 0.116, p= 2.6E-04) and the ΔDD (β=0.242 95%CI: 0.097 to 0.388, p= 1.5E-03). That is, for each day that the ΔDD increases, the Td10 increases by 0.242 days and for each day that passes from the diagnosis of the first case, the Td10 increases 0.077 days. These two variables explain 29.7% of the variability of the Td10 (r^2^= 0.297) (Table 1). It is not rare that temperature stands out of the model, as there is a very high correlation between temperature and ascending dates in the calendar from January to March. In fact, when the date of the first case is not introduced into the MLR model, the temperature remains significant in the model (data not shown). However, the exclusion of the temperature in the model and the permanence of the date of diagnosis of the first case may have an additional explanation.

**Table 1.** Liner regression models to evaluate explanatory variables of doubling time the first and second 10 days of the growth curve of Covid-19 in 59 countries.

| Variables<sup>b</sup> | Lineal regression models<sup>a</sup> |  |  |  |  |  |
| --- | --- | --- | --- | --- | --- | --- |
|  | Univariate |  |  | Multivariate |  |  |
|  | β (95% CI) | p-value | r<sup>2</sup> | β (95% CI) | p-value | r<sup>2</sup> |
| Td10 (n=59) |  |  |  |  |  |  |
| DCS | -0.072 (-0.146-0.002) | 5.8E-02 | .044 | 0.007 (-0.083-0.097) | 8.8E-01 | 0.297 |
| D100 | -0.019 (-0.089-0.05) | 5.8E-01 | 0 |  |  |  |
| ΔDD | 0.243 (0.081-0.406) | 4.1E-03 | .121 | 0.242 (0.097-0.388) | 1.5E-03 |  |
| Date first case | 0.079 (0.037-0.121) | 4.0E-04 | .182 | 0.077 (0.037-0.116) | 2.6E-04 |  |
| Average Relative humidity | -0.012 (-0.079-0.056) | 7.3E-01 | 0 |  |  |  |
| Average temperature | 0.17 (0.092-0.247) | 4.7E-05 | .237 | 0.064 (-0.044-0.173) | 2.4E-01 |  |
| Climtic zone | 2.86 (1.191-4.529) | 1.1E-03 | .154 | -0.663 (-3.359-2.034) | 6.2E-01 |  |
| Td20 (n=45) |  |  |  |  |  |  |
| Td10 | 0.793 (0.291-1.295) | 2.7E-03 | .172 | 0.999 (0.468-1.53) | 4.9E-04 | 0.426 |
| DCS | -0.087 (-0.184-0.011) | 8.2E-02 | .047 | 0.322 (0.075-0.57) | 1.2E-02 |  |
| D100 | -0.071 (-0.16-0.017) | 1.1E-01 | 0 | -0.332 (-0.549--0.115) | 3.7E-03 |  |
| ΔDD | -0.009 (-0.243-0.226) | 9.4E-01 | 0 |  |  |  |
| Date first case | 0.103 (0.024-0.181) | 1.1E-02 | .120 | 0.077 (-0.028-0.182) | 1.5E-01 |  |
| Average Relative humidity | -0.027 (-0.123-0.069) | 5.7E-01 | 0 |  |  |  |
| Average temperature | 0.151 (0.018-0.284) | 2.7E-02 | .088 | -0.161 (-0.345-0.023) | 8.4E-02 |  |
| Climtic zone | 4.022 (1.877-6.167) | 4.8E-04 | .232 | 5.704 (2.396-9.012) | 1.2E-03 |  |

a. The variables that pass in the univariate models at $p < 0.2$ were introduced in the multivariate models using the backward method.
b. Td10 and Td20 = doubling time the first 10 and 20 days of the curve, respectively, DCS= day start the curve climb, D100= day when the 100th was diagnosed, ΔDD=D100-DCS, climatic zones = 1 for temperate zones and 2 for tropic/subtropic zones in the models.

In the ULR models for Td20, all variables are significant, except relative humidity and ΔDD. But only four variables remained in the MLR model, including Td10 (β=0.999 CI95%: 0.468 to 1.53, p= 4.91E-04) and the climate zone (β=5.7 CI95%: 2.396 to 9.012, p= 1.22E-03), which appeared to be the most important factors influencing the value of Td20 (Table 1). For each day that the Td10 increases, the Td20 increases by 0.999 days, conversely, for each day the Td10 decreases, the Td20 will decrease by 0.999 days. In tropical/subtropical countries, Td20 increases 5.7 days above the Td20 of temperate countries. For Td20, the temperature no longer seems to be an important factor, while important components of the initial curve behavior remained, such as DCS and D100 (Table 1). All these factors explain 42.6% (r^2^ = 0.426) the variability of Td20. When the model is stratified by zone, the Td10 variable remains in both area models as the most important variable (data not shown).

### Prediction of total cases of Covid-19 at days 20 and 30 of the growth curve: MLR models

We explored whether the Td10, Td20, and TdT, with the other variables, could predict the Covid-19 CF that would be reached 20 and 30 days after the day D100 in the growth curve using ULR and MLR models. Variables that had a p=0.2 in the ULR models were selected to be introduced into the MLR model. A clear relationship of Td10 with CF at day 20 is observed in the ULR model (r^2^ = 0.189), as discussed above in relation to Figures 4 and 6, however, the relationship of TdT to the CF is much higher (r^2^= 0.268), therefore this variable of the doubling time was the one introduced in the MLR model (Table 2). Also, ΔDD, date of first case, the average relative humidity, the average temperature and climate zone passed the cut-off value in the ULR models (Table 2). However, in the MLR analysis only TdT, date of first case and the average temperature remained in the model. These three variables had a negative relationship with the CF, that is, as their value increases the CF decreases, and vice versa, when their value decreases, the CF increases. The explanatory variables had a linear correlation with the CF (r= -0.69) and explain 44.7% of the variability of CF for day 20 of the curve. This indicates that other factors, such as epidemic containment measures, would explain more than 55% of the remaining variability.

**Table 2.** Prediction of the accumulative frequency of Covid-19 at day 20 and 30 of the curve with the doubling time and other variables using linear regression models.

| Variables<sup>b</sup> | Univariate regression models<sup>a</sup> |  |  | Multivariate regression models |  |  |
| --- | --- | --- | --- | --- | --- | --- |
|  | β (95% CI) | p-value | r<sup>2</sup> | β (95% CI) | p-value | r<sup>2</sup> |
| Prediction of total cases of Covid-19 at day 20 of the curve* |  |  |  |  |  |  |
| Td10 | -1590.9 (-2729.3 - -452.6) | 7.5E-03 | .182 |  |  | 0.449 |
| Td20 | -978.6 (-1646.5 - -310.7) | 5.3E-03 | .197 |  |  |  |
| TdT | **-2381.4 (-3713 - -1049.8)** | **8.8E-04** | **.268** | **-1912.9 (-3161.6--664.2)** | **3.7E-03** |  |
| FC<sub>Td20/Td10</sub> | -557.2 (-3068.1-1953.7) | 6.6E-01 | 0 |  |  |  |
| DCS | 27.9 (-199.9-255.8) | 8.1E-01 | 0 |  |  |  |
| D100 | -30.8 (-233.1-171.5) | 7.6E-01 | 0 |  |  |  |
| ΔDD | **-374.4 (-898.5-149.7)** | **1.6E-01** | 0 | -219.6 (-700-260.8) | 3.6E-01 |  |
| Date of first case | **-252.3 (-449.4--55.3)** | **1.4E-02** | **.158** | **-164 (-342.4-14.3)** | **7.0E-02** |  |
| Average Relative humidity | 93.5 (-160.1-347.1) | 4.6E-01 | 0 |  |  |  |
| Average temperature | **-477.9 (-764.2 - -191.7)** | **1.7E-03** | **.242** | **-259.9 (-545-25.2)** | **7.3E-02** |  |
| Climtic zone | **-8612.2 (-13559 - -3665.5)** | **1.2E-03** | **.257** | -1395.8 (-9320-6528.3) | 7.2E-01 |  |
| Prediction of total cases of Covid-19 at day 30 of the curve** |  |  |  |  |  |  |
| Td10 | -5003.8 (-13121-3113.5) | 2.1E-01 | 0 |  |  | 0.638 |
| Td20 | **-14255.8 (-23467.2--5044.4)** | **4.7E-03** | **.365** | **-17576.1 (-27101.2--8051)** | **1.7E-03** |  |
| Td30 | -1008.6 (-2691.2-674.1) | 2.2E-01 | 0 |  |  |  |
| TdT | -15583.5 (-26521.1--4645.9) | 8.1E-03 | .323 |  |  |  |
| FC<sub>Td20/Td10</sub> | -10862.8 (-35352.9-13627.4) | 3.6E-01 | 0 |  |  |  |
| FC<sub>Td30/Td20</sub> | -5028.7 (-21031.3-10974) | 5.1E-01 | 0 |  |  |  |
| DCS | 682.9 (-1207-2572.7) | 4.5E-01 | 0 |  |  |  |
| D100 | 541.6 (-1225.4-2308.4) | 5.3E-01 | 0 |  |  |  |
| ΔDD | -239.6 (-4176.2-3697) | 9.0E-01 | 0 |  |  |  |
| Date of first case | **-1437.3 (-4068.5-1193.9)** | **2.6E-01** | 0 | **-2537.5 (-4556.9--518)** | **1.8E-02** |  |
| Average Relative humidity | -634.2 (-3049.7-1781.2) | 5.9E-01 | 0 |  |  |  |
| Average temperature | **-1844.2 (-4980.7-1292.3)** | **2.3E-01** | 0 | -3299.4 (-8736.1-2137.2) | 2.1E-01 |  |

a. Univariate and multivariate models using the ENTER method.
b. Td10 and Td20 = doubling time the first 10 and 20 days of the curve, respectively, DCS= day start the curve climb, D100= day when the 100th was diagnosed, ADD=D100-DCS, climatic zones = 1 for temperate zones and 2 for tropic/subtropic zones in the models.
\*The parameters of the multivariate model: b=23,067, r=0.67, Durbin-Watson=1.65, ANOVA p<0.0001
\*\* The parameters of the multivariate model: b=161,049, r= 0.8, Durbin-Watson=1.9, ANOVA p=0.002

For the prediction of the CF at the 30th day of growth curve the model has a better performance (Table 2), but was explored only for temperate areas, since in tropical/subtropical areas very few countries of those analyzed in the current study had reached 30 days of evolution in the curve at the time of writing this article. The Td20 performed better than the TdT, so it was this variable that was introduced in the MLR model. In fact, Td20 alone explains 36.5% of the CF’s variability at the 30th day of the growth curve. This indicates that the speed of the growth curve during the previous 10 days (days 11 to 20), is essential for controlling the CF during the following 10-day period of the outbreak evolution, that is, from 21 to 30 days after the day D100. Since none of the other factors analyzed with the ULR models passed the cut-off value, in addition to Td20, the date of the first case and the average temperature were introduced into the MLR analysis since the p-values are just above 0.2. In the MLR analysis, only Td20 and the date of the first case remained in the model, which explained 63.8% (r^2^= 0.638) of the variability of the frequency of accumulated infected cases towards the 30th day of the curve. Similarly, both factors have a negative influence on the CF. The rest of the variability (36%) must be related to other factors, such as the containment measures used by each country to control the epidemic.

## Discussion

### Key results

In this study we disclosed that the behavior of the growth curve during the period between the diagnosis of the initial and the first 100 cases, especially the day the curve begin to rise (DCS), the day the first 100 cases (D100) were reached and the difference between these two time points (ΔDD), were related to temperature, the date of the first case and the doubling time (Td). In addition, these values were substantially different between countries located in temperate and tropical/subtropical areas, especially the Td during the first ten days (Td10), after D100, was on average 3 days longer in tropical/subtropical than in temperate countries. We also identified that the factors involved in Td the first ten (Td10) days (date of first case diagnosed and ΔDD) are different than those involved the second ten (Td20) days (mainly Td10 and climate zone) of the growth curve. The fastest growth curves during the first 10 days, after D100 day, were associated with Td10 <2 and Td10< 3 in temperate and tropical/subtropical countries, respectively. And the fold change Td20/Td10 > 2 was associated with earlier flattening of the growth curve. In the MLR predictive models, the Td10, the date of the first Covid-19 case diagnosis and the ambient temperature, were negatively related to the cumulated frequency for the 20^th^ day of the growth curve, while only Td20 and date of the first Covid-19 case were negatively related to the CF reached on day 30 of the growth curve, although for the latter, only the data of temperate countries was used.

### Comparison with previous studies and possible explanations of our findings

Considering the fact that the Td was calculated based on the CF reported daily by the WHO, there is great uncertainty about the variability of this information with the reality in each country depending on the number of tests carried out in each of them. The number of tests have been changing and we do not have a reliable information about the time the mass use of SARS-Cov-19 screenings began in each country, nor how many tests were performed by days 10, 20 and 30 days of the growth curve, after the day that case 100 (D100) was reached. Moreover, the promptness and type of containment measures adopted by each country could have also affected the Tds. However, we can discuss which data from our analysis could have little or a lot of deviation related to the number of tests and containment measures, and therefore the importance of the findings.

Notably, the findings from the analysis of the first stage of the growth curve, from the day on which the first case was diagnosed until the first 100 cases of Covid-19 were diagnosed, as well as the date on which the first diagnosis was made and probably the value of Td10, would have a modest influence from the number of tests or containment measures that were carried out in the different countries, since most of them were similar. The date of the first case diagnosed, the average differences in the day on which the growth curve raised (DCS) and from there to the time of the first 100 cases (ΔDD) were significantly different among temperate and tropical/subtropical countries. Although the date of the first case diagnosed is directly related to temperature, this variable remains independent and heavier in the MLR models associated to Td10 and CF. The above suggest that the date of the first case is not only related to ambient temperature but perhaps also related to the way the infection was disseminated. The fact that the epidemic reached the tropical/subtropical countries at a later time could have been an explanation as a delay in the shockwave from China, first to other temperate countries in the northern hemisphere and from there to tropical/subtropical countries. For example, the infection in European countries and the USA, took an estimate, between the number of days for the rise of the curve and an important establishment of the infection, of about 2 months to be exported to Latin America and other countries of tropical/subtropical areas. The average time delay, relative to December 29^th^, 2019, when the infection spread from China, was 58.5 ± 15.2 days. The question is why in these countries the cumulated positive cases curve rose rapidly but with a smaller slope than in countries of temperate zones, and why in the later ones, once the rise of the curve began, the slope was steeper. In discussing the last issue first, an important possibility could be that in European and other temperate countries, asymptomatic individuals were the first to accumulate and these subjects spread the infection.^19^ This seems very feasible as up to 80% of infected individuals are asymptomatic.^20, 21^ 22, 23 In addition, common use of mass transportation and small housing spaces in European cities or those such as New York with a high population density, could have facilitated infection dissemination. Further, the spread could have then affected the older population which in temperate countries make up a much higher percentage of the population pyramid than in countries of tropical/subtropical areas. For example, in countries like Spain and Italy the 65 years and older age-group, constitutes 19% and 23% of their populations, respectively. Conversely, the same age-group represents only 7% of the Mexican and Ecuadorean populations. The older population may have greater susceptibility to infection in cold climates, and it has been suggested that social and biological factors may interact. For example, factors such as cohabitation in small spaces with poor ventilation^10^ may interact with a decreased immune response associated with aging, and diminished airway function^,24-27^ further predisposing to viral infections. This would explain the late rise with a steeper slope in the epidemic curve in those countries. In contrast, in tropical/subtropical countries the early ascent, with a slower slope of the curve, could be related to the initial presence of imported cases, mainly from European countries and the US, and the dissemination to their contacts. We have hypothesized that these imported cases, mainly of middle or higher socioeconomic groups who have the possibility of traveling both for leisure and work, could have later spread the infection on a smaller scale, mainly because they do not use mass public transportation. On the other hand, in the population pyramid in those countries there is a large proportion of young people, which could have contributed to a lower frequency rate, due to a silent outbreak or an infection with less severe clinical manifestations. For example, the median age of confirmed cases with Covid- 19 is much lower in Mexico, according to the daily data reported by the Secretary of Health of this country (45 years old),^28^ than that in China (56 years old)^29^ and Italy (64 years old).^30^

In addition, the expectation in these tropical/subtropical countries, is that there is a protective effect of ambient temperature, at least on the airway function.^8^ Temperature variation as a function of climatic status has a profound influence in virtually all stages of the host virus interaction;^31-33^ therefore, the relationship of ambient temperature and doubling time depicted here becomes highly relevant. There is a direct effect of temperature on the enzyme-mediated reactions resulting in cell homeostasis and immune responses.^31, 34^ More explicitly, the frequency of upper respiratory tract viral infections relates inversely to every degree Celsius that the temperature drops.^31, 33, 34^ Although available information strongly suggests a climatic influence on Covid-19 infection and spreading,^18, 32^ the novelty of this illness, warrants further assessment of this issue. While the information gathered herein provides further support to the notion that an overall increased temperature and humidity limits the spreading and occurrence of Covid-19, systematic studies are required to ruled out possible ethnical and/or genetical influences^35^ upon the spreading of and susceptibility to the disease. In this regard, as mentioned earlier, the human ACE2 receptor has now been recognized as the receptor for the SARS-CoV-2 S protein, and variations in this gene may confer susceptibility or some type of resistance to the viral infection.^36^ Allelic variants may differ according to countries or populations and explain, at least partially, differences in infection rates. On the other hand, the differences in the rise of the curve, the ΔDD and the Td10 were not uniform among the countries within temperate or tropical/subtropical areas. For example, in European countries such as Switzerland, Austria, Holland, Norway and the Czech Republic, these parameters were similar to those observed in tropical/subtropical areas and not to those in the rest of Europe. This finding coincides with the diagnosis of Covid-19 later than in the rest of Europe (Figure 3 panel A). In fact, these countries and the tropical/subtropical ones had more time to implement containment measures, which could have also contributed to the increase of Td.

In the case of Td20, there is a possibility that this calculation is biased or at least severely related to the number of tests and containment measures that each country has undertaken and may have contributed, at least in part, to the differences between temperate and tropical/subtropical countries. If this would have been the case, we would have observed a greater difference between the comparisons of partial doubling times between the climate zone as the curve progresses, that is, that there would have been a greater difference (Δ) in ΔTd30 >ΔTd20 > ΔTd10. We were not able to compare the Td30 as the curve in most of the countries in tropical/subtropical areas have not reached that period yet, but we found that ΔTd20>ΔTd10. Although, in the lineal regression models, Td10 participated in the value of Td20, only explaining about 17% of the variability of that value. Therefore, the expected difference in the Td20s between countries within temperate and tropical/subtropical zones should not be greater than 1.17 times Td10, and in opposition to that, the difference of the Td20s was 2.6 times that of Td10. This suggests that the differences in Td20 between countries in the different climate areas, may be related to factors such as the number of tests performed and the implemented measures to mitigate the epidemic. However, the relationship or change folds between Td20 and Td10 must remain valid, on the assumption, at least in principle, that the policies on the number of tests and implemented containment measures, did not change too much in each country. In addition, no statistically significant difference was found between FC_Td20/Td10_ in countries within temperate or tropical/subtropical regions. This implies that the value of FC_Td20/Td10_ ≥ 2 would be a good parameter for both regions to positively assess the evolution of the epidemic during the 21-30 day-periods of the curves.

## Strengths and limitations

This study has several strengths including the large sample of incident cases of Covid-19 collected from the WHO database, to analyze the behavior of the epidemic curves for more than 90 days. Another strength is the fact that epidemic curves and environmental variables of 67 countries distributed in different climatic zones and six continents, were studied. These two issues allowed the comparison of several variables in various segments of the epidemic growth curve and to establish the differences between the two climatic zones. Notwithstanding, the study has some limitations such as the inability to incorporate in the analysis the number of tests carried out or the containment measures implemented in each country, during the different periods of the epidemic curves. Although it is possible that these issues most likely do not affect or have very little impact on the analysis during the first 10 days of the epidemic curve, it is likely that they substantially affect the analysis in the second ten and third ten days of the epidemic curves. Despite this, the fold change ratio between Td20 and Td10 was no different between the two compared climate zones.

## Implications

Having a different pattern of infection spread between temperate and tropical/subtropical countries could only slow the speed in which the virus is being transmitted, and although this is good news for health services utilization, it does not necessarily imply that the proportion of the population infected will be smaller in the tropical/subtropical countries.^9^ The latter will depend on the timing, related to the cumulative frequency growth curve, in which the containment measures are established, and their magnitude to reduce the spread of the epidemic in each country.^9, 37-39^ On the other hand, the value of Td10 allows the evaluation of the growth of infections during the second 10 days of the epidemic curve, while the ratio Td20/Td10 allows the study to evaluate the growth of the curve during the third 10 days and with it, may help in evaluating how effective are the implemented containment measures.

## Conclusions

This study showed that the behavior of the growth curve (DCS, D100 and ΔDD) in the first stage of the epidemic is related to the date that the first Covid-19 case was diagnosed, the ambient temperature and doubling time of infection cases, which are different between countries located in temperate and tropical/subtropical areas. The Td10 is on average 3 days longer in tropical/subtropical countries than in those located in temperate zones and can predict the growth of the curve for the following 10 days of the evolution of the epidemic after D100. These differences appear to be related to ambient temperature and the date of the first case that was identified and how the infection spread in both climatic zones. In addition, the Td10 and Td20 values helped predict the cumulative frequency of Covid-19 at the 20^th^ and 30^th^ days of the epidemic after D100 day, while the Td20/Td10 ratio helps to evaluate the growth curve behavior during the next third 10 days of the epidemic evolution.

## Data Availability

All data generated or analysed during this study are included in this article.

## What is already known on this topic

- Data from China and other Asian countries during the first month of the Covid-19 pandemic suggest that ambient temperature and humidity have a direct relationship to the doubling time (Td) for the number of cases.

## What this study adds

- There is a relationship not only between the ambient temperature, but also with the date when the first case was diagnosed and the behavior of the growth curve of the cumulated frequency, with the doubling time during the different 10-day time periods of the pandemic growth in 67 countries around the world.
- Very clear differences exist in the time of duplication (Td) of cases among countries located in temperate and tropic/subtropical climate areas. Also, it calculates how much of the variability of the cumulated frequency of Covid-19 is explained by the Td and other environmental variables.

